# Limited specificity of serologic tests for SARS-CoV-2 antibody detection, Benin, Western Africa

**DOI:** 10.1101/2020.06.29.20140749

**Authors:** Anges Yadouleton, Anna-Lena Sander, Andres Moreira-Soto, Carine Tchibozo, Gildas Hounkanrin, Yvette Badou, Carlo Fischer, Nina Krause, Petas Akogbeto, Edmilson F. de Oliveira Filho, Anges Dossou, Sebastian Brünink, Melchior A. Joël AÏssi, Mamoudou Harouna Djingarey, Benjamin Hounkpatin, Michael Nagel, Jan Felix Drexler

## Abstract

Testing 68 RT-PCR-confirmed COVID-19 cases and controls from Benin, Western Africa with commercially available SARS-CoV-2 antibody ELISAs revealed up to 25% false-positive results, likely due to unspecific antibody responses elicited by acute malaria. Serologic tests must be carefully evaluated to robustly assess SARS-CoV-2 spread and immunity in tropical regions.

## Introduction

Since its emergence in China late 2019, Coronavirus disease-19 (COVID-19) has afforded over 25 million cases and over 850,000 deaths globally by September 2020. Diagnosis of the causative pathogen severe acute respiratory syndrome-coronavirus-2 (SARS-CoV-2) is commonly based on reverse-transcription polymerase chain reaction (RT-PCR) detecting viral nucleic acid or serologic assays based on antigen detection in early stages of disease (1, 2). In later stages of disease, antibody-based serologic testing can complement diagnosis of SARS-CoV-2 infection. In addition, antibody-based serologic testing is a valuable epidemiological tool to assess spread and potential immunity to SARS-CoV-2. Serologic studies in European and Asian countries indicate high sensitivity and specificity of widely used SARS-CoV-2 antibody ELISAs (3, 4). However, many serological tests have not been validated in resource-limited settings (5). For this purpose, we conducted a SARS-CoV-2 serologic assessment using SARS-CoV-2 RT-PCR-confirmed patients and controls in Benin, Western Africa.

## The study

We obtained convalescent sera from eight RT-PCR-confirmed patients sampled during March-April 2020 immediately after the identification of the first COVID-19 cases in Benin (average sampling was 8 days post SARS-CoV-2 RT-PCR confirmation; range: 1-10 days; **Table 1**) and 60 sera from patients with acute febrile illness sampled for hemorrhagic fever surveillance during October-November 2019 (**Table 2**; sampling approved by the ethics committee of the Benin Ministry of Health: Arrêté 2020 No. 030/MS/DC/SGM/DNSP/CJ/SA/027SGG2020).

**Table 1:**
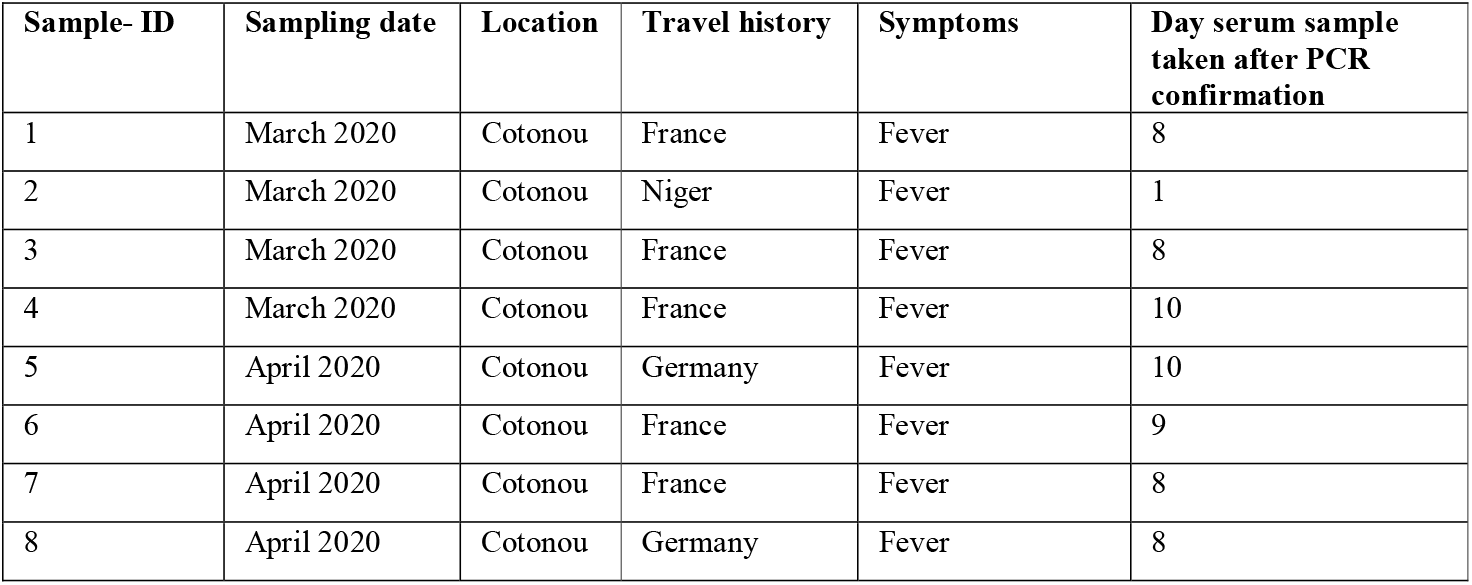
Characteristics of confirmed SARS-CoV-2 positive patients for which serum samples were available taken from March-April 2020

| Sample- ID | Sampling date | Location | Travel history | Symptoms | Day serum sample taken after PCR confirmation |
| --- | --- | --- | --- | --- | --- |
| 1 | March 2020 | Cotonou | France | Fever | 8 |
| 2 | March 2020 | Cotonou | Niger | Fever | 1 |
| 3 | March 2020 | Cotonou | France | Fever | 8 |
| 4 | March 2020 | Cotonou | France | Fever | 10 |
| 5 | April 2020 | Cotonou | Germany | Fever | 10 |
| 6 | April 2020 | Cotonou | France | Fever | 9 |
| 7 | April 2020 | Cotonou | France | Fever | 8 |
| 8 | April 2020 | Cotonou | Germany | Fever | 8 |

**Table 2:** Characteristics of patients with febrile illnesses of unknown origin taken from October-November 2019

| Sample- ID | Health Center* | Sampling date | Symptoms |
| --- | --- | --- | --- |
| 215 | CNHU | October 2019 | Fever |
| 311 | CB | October 2019 | Fever |
| 312 | CB | October 2019 | Fever |
| 313 | CB | October 2019 | Fever |
| 314 | CB | October 2019 | Fever |
| 315 | CB | October 2019 | Fever |
| 316 | CB | October 2019 | Fever |
| 317 | CB | October 2019 | Fever |
| 318 | CB | October 2019 | Fever |
| 319 | CB | October 2019 | Fever |
| 320 | CB | October 2019 | Fever |
| 321 | CB | October 2019 | Fever |
| 322 | CB | October 2019 | Fever |
| 323 | CB | October 2019 | Fever |
| 324 | CB | October 2019 | Fever |
| 325 | CB | October 2019 | Fever |
| 326 | CB | October 2019 | Fever |
| 327 | CB | October 2019 | Fever |
| 328 | CB | October 2019 | Fever |
| 329 | CB | October 2019 | Fever |
| 330 | CB | October 2019 | Fever |
| 331 | CB | October 2019 | Fever |
| 332 | CB | October 2019 | Fever |
| 333 | CB | October 2019 | Fever |
| 334 | CB | October 2019 | Fever |
| 335 | CB | October 2019 | Fever |
| 336 | CB | October 2019 | Fever |
| 337 | CB | October 2019 | Fever |
| 338 | CB | October 2019 | Fever |
| 339 | CB | October 2019 | Fever |
| 201 | CB | November 2019 | Fever |
| 202 | CB | November 2019 | Fever |
| 203 | CB | November 2019 | Fever |
| 204 | CB | November 2019 | Fever |
| 205 | AHC | November 2019 | Fever |
| 206 | AHC | November 2019 | Fever |
| 207 | AHC | November 2019 | Fever |
| 208 | AHC | November 2019 | Fever |
| 209 | AHC | November 2019 | Fever |
| 210 | AHC | November 2019 | Fever |
| 211 | AHC | November 2019 | Fever |
| 212 | AHC | November 2019 | Fever |
| 213 | AHC | November 2019 | Fever |
| 214 | AHC | November 2019 | Fever |
| 216 | CNHU | November 2019 | Fever |
| 217 | CNHU | November 2019 | Fever |
| 218 | CNHU | November 2019 | Fever |
| 219 | CNHU | November 2019 | Fever |
| 220 | CNHU | November 2019 | Fever |
| 221 | CNHU | November 2019 | Fever |
| 222 | CNHU | November 2019 | Fever |
| 223 | CNHU | November 2019 | Fever |
| 224 | CNHU | November 2019 | Fever |
| 225 | CNHU | November 2019 | Fever |
| 226 | CNHU | November 2019 | Fever |
| 227 | CNHU | November 2019 | Fever |
| 228 | CNHU | November 2019 | Fever |
| 229 | CNHU | November 2019 | Fever |
| 230 | CNHU | November 2019 | Fever |
| 291 | AHC | November 2019 | Fever |

\* Samples originated from three major health centers: Akkasato Health Center (AHC), Centre National Hospitalier Universitaire Hubert Koutoukou MAGA (CNHU) and the Clinique Boni (CB).

Sera were tested using commercially available ELISAs relying on different antigens and antibody classes, namely SARS-CoV-2 nucleocapsid (N) antigen (IgG), spike S1 subunit (both IgG and IgA), Middle East respiratory syndrome coronavirus (MERS)-CoV S1 (IgG; all from Euroimmun, Germany), and the FDA-approved SCoV-2 detect SARS-CoV-2 ELISA (IgG only; spyke antigen; InBios, USA). Additionally, sera were tested using commercially available ELISA kits (Euroimmun, Germany) against the Zika virus (ZIKV) NS1 antigen (IgG), the Epstein-Barr virus (EBV) EBNA1 antigen (IgG), the EBV VCA antigen (both IgM and IgG) and using real time-PCR for Plasmodia (all human pathogenic species), EBV and cytomegalovirus (CMV) (all PCR tests from TIB Molbiol, Germany). Plaque reduction neutralization tests (PRNT) were performed using similar methods for SARS-CoV-2 and for ZIKV as previously described (4, 7). Testing for common cold betacoronaviruses HCoV-OC43- and HCoV-HKU1-specific antibodies relied on recombinant spike protein-based immunofluorescence assays as previously described (8).

In the eight RT-PCR-confirmed patients, SARS-CoV-2 seroconversion ranged from 62.5 to 100% (8/8; 95% CI: 30.8-100.0%), depending on the ELISA that was used (**Figure 1A**). This observation suggested differential sensitivity of ELISAs based on the immunoglobulin detected and on the commercial kit used. Indeed, IgA-based had a higher sensitivity compared to most of IgG-based SARS-CoV-2 ELISAs early after infection and only the InBios IgG-based kit detected all RT-PCR confirmed patients as positive (**Figure 1A**) (4). As shown in **Figure 1B**, 87.5% (7/8) of those ELISA results were confirmed by a highly specific SARS-CoV-2 PRNT. In 60 samples taken during October-November 2019 from febrile patients, 25.0% positive or borderline ELISA results that potentially represent true positives were observed when summarizing all antibody classes, antigens and kits (15/60; 95% CI, 15.7-37.3) (9). Different from RT-PCR-confirmed cases, ELISA reactivity in those samples contrasted with the complete lack of SARS-CoV-2-specific neutralizing antibodies (**Figure 1A** and **1B**). Likely unspecific SARS-CoV-2 ELISA reactivity may be consistent with three scenarios. First, antibodies elicited by common infections with endemic human coronaviruses may cross-react with SARS-CoV-2 antigens in patients (1). However, sera that yielded positive SARS-CoV-2 ELISA results did not differ significantly from sera that yielded negative SARS-CoV-2 ELISA results in their reactivity with common cold coronaviruses (45.7-63.6% versus 70.4-74.0%; p=0.1 and p=0.7, Fisher’s exact test) (**Figure 1C**). Similarly, the magnitude of antibody titers against common cold coronaviruses did not differ significantly between those groups (p=0.09 and p=0.8, t-test) (**Figure 1D**). Notably, no serum reacted with MERS-CoV antigens, suggesting that unspecific reactivity may not automatically apply to all coronavirus antigens and tests (**Figure 1E**). Second, polyclonal B-cell activation can occur in infections with or reactivations of herpesviruses such as CMV and EBV and elicit false-positive results in serologic tests (10). However, only two patients were positive in a CMV PCR and only one patient was positive in an EBV PCR (**Figure 2A**). Additionally, SARS-CoV-2 ELISA-positive versus ELISA-negative individuals did not differ in their past exposure to EBV according to detailed serologic analyses (**Figure 2A** and **2B)**. Lastly, polyclonal B-cell activation can also be caused by acute malaria, which is widespread in Africa (11). As shown in **Figure 2C**, a higher proportion of those individuals that yielded positive SARS-CoV-2 ELISA results than those that yielded negative ELISA results were positive for Plasmodia in a highly sensitive PCR test (71.4% versus 54.3%), albeit this difference was not statistically significant (p=0.35, Fisher’s exact test). Similarly, significantly higher parasite loads occurred within SARS-CoV-2 ELISA-positive compared to ELISA-negative individuals (**Figure 2C**) (p=0.035; t-test). Higher parasite loads that decrease overtime have been observed in acute malaria, suggesting a higher proportion of acute malaria in SARS-CoV-2 ELISA-positive patients compared to sub-acute or chronic malaria in SARS-CoV-2 ELISA-negative patients (12). To assess the breadth of potentially unspecific reactivity, we tested the sera from febrile patients using a ZIKV IgG-ELISA for which unspecific reactivity in cases of acute malaria has been reported previously (11). As shown in **Figure 2D**, sera that elicited potentially unspecific SARS-CoV-2 ELISA results also elicited significantly more frequently positive ZIKV ELISA results (57.1 versus 23.9%; p=0.019, Fisher’s exact test). None of the sera yielding positive ZIKV ELISA results showed ZIKV-specific neutralizing antibodies, suggesting unspecific reactivity of those sera in the ZIKV ELISA (**Figure 2A; Figure 2E**). Additionally, sera that yielded potentially false-positive results in the SARS-CoV-2 ELISA were also significantly more likely to show potentially false-positive results in the ZIKV ELISA (p=0.04; Chi-Square test) (**Figure 2D** and **2F**).

**Figure 1.**
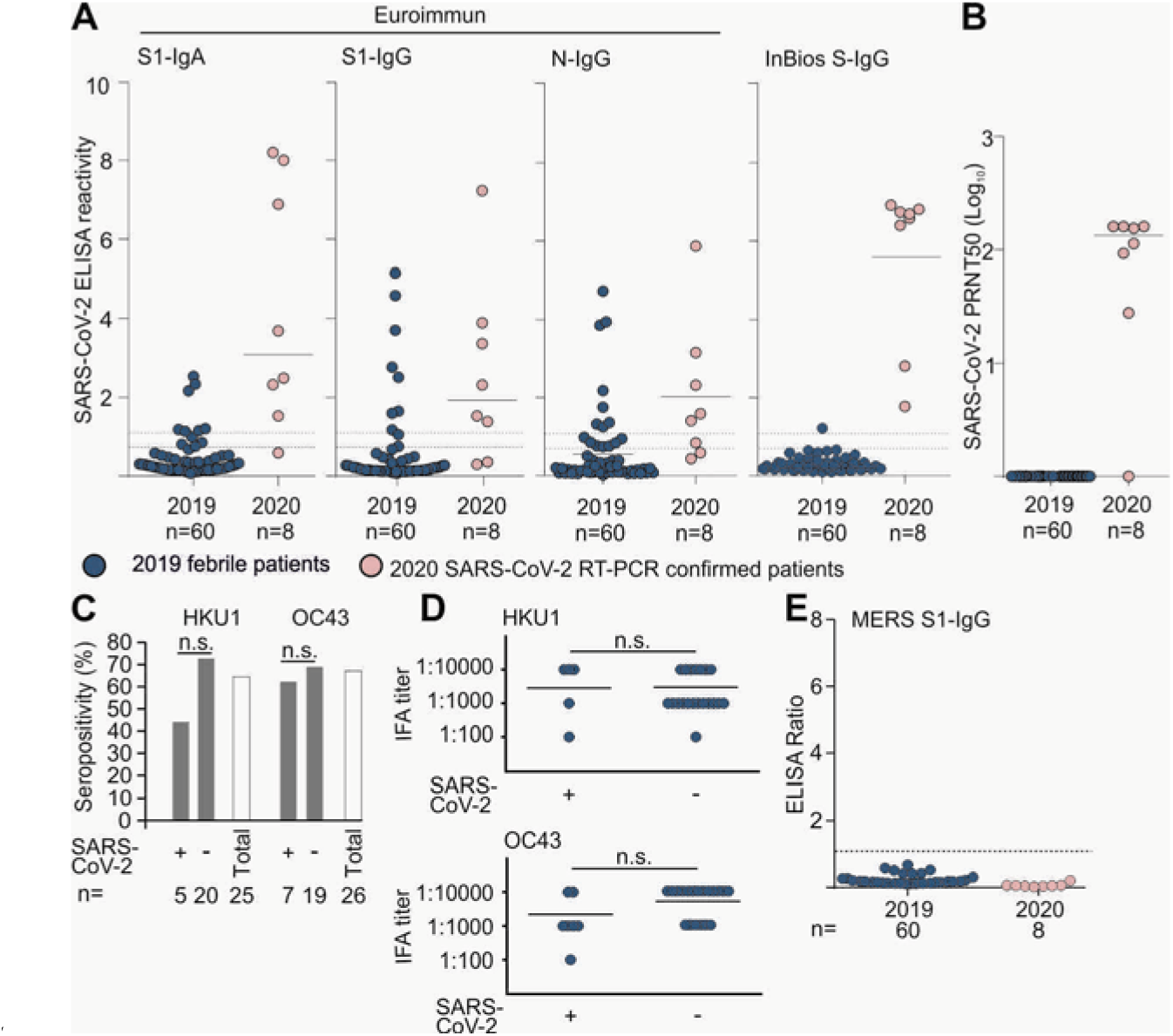
Serologic diagnostics of betacoronaviruses in Benin. **A**) SARS-CoV-2 ELISA reactivity using different commercially available assays in febrile patients from 2019 and SARS-CoV-2 RT-PCR-confirmed patients from 2020. Dashed lines denote the ratio positivity threshold of >1,1 and borderline results between >0.9 to <1.1 defined by the manufacturer. Continuous line denotes the mean ELISA reactivity. **B**) SARS-CoV-2 plaque reduction neutralization test (PRNT_50_) in febrile patients from 2019 and SARS-CoV-2 RT-PCR-confirmed patients from 2020, shown in Log_10_ scale for clarity of presentation. Continuous line denotes the mean PRNT Log_10_ titer. **C**) Common cold betacoronaviruses HKU1 and OC43 seropositivity between 2019 febrile patients that were SARS-CoV-2 ELISA positive versus SARS-CoV-2 ELISA negative. **D**) Common cold betacoronaviruses IFA titer in febrile patients from 2019 and SARS-CoV-2 RT-PCR-confirmed patients from 2020. Samples that were negative are not shown for graphicalreasons. N.s. not significant. **E**) MERS-CoV ELISA ratio in febrile patients from 2019 and SARS-CoV-2 RT-PCR confirmed patients from 2020. Dashed line denotes the ratio positivity threshold of >1.1 ratio defined by the manufacturer.

**Figure 2.**
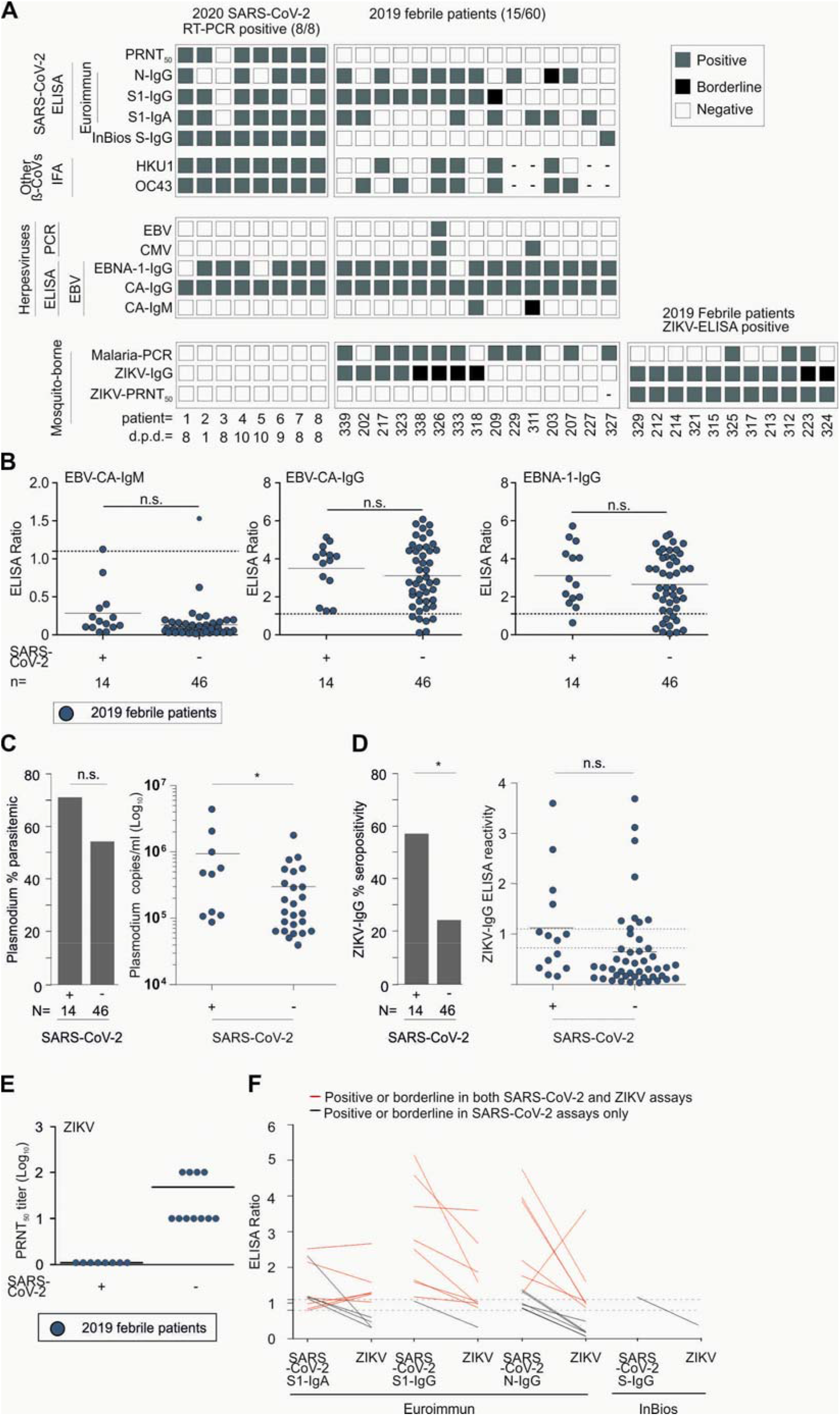
Molecular and serologic test results for endemic pathogens in Benin. **A**) Individual reactivity of different commercially available SARS-CoV-2 ELISAs, SARS-CoV-2 plaque reduction neutralization test (PRNT) and Immunofluorescence (IFA) reactivity to common cold betacoronaviruses (ß-CoVs) OC43 and HKU1 in febrile patients from 2019 and SARS-CoV-2 RT-PCR confirmed patients from 2020. Individual results of Epstein-Barr-Virus (EBV)-PCR, Cytomegalovirus (CMV)-PCR and three EBV ELISA: EBV-IgM, EBV-IgG and EBNA-IgG from the same patients. Individual results of Zika virus (ZIKV)-IgG ELISA, ZIKV-PRNT and malaria-PCR from the same patients. Gray square denotes a positive result, black square denotes an inconclusive result and white squares a negative result. Samples in which the assay was not performed due to low sample volumes are marked with a hyphen (-). d.p.d. denotes days the serum sample was taken post positive RT-PCR SARS-CoV-2 diagnosis. **B**) Epstein-Barr virus (EBV)-IgM, EBV-IgG and EBV-EBNA-IgG ELISA ratio between 2019 febrile patients that were SARS-CoV-2 ELISA positive versus SARS-CoV-2 ELISA negative. Dashed lines denote the ratio positivity threshold defined by the manufacturer. Continuous line denotes the mean ELISA reactivity. N.s. not significant. **C**) Percent parasitemic febrile patients that were SARS-CoV-2 ELISA-positive versus SARS-CoV-2 ELISA-negative. N.s. not significant. Right: Log_10_ plasmodium copies per ml. Continuous line denotes the mean copies/ml. Asterisk denotes p<0.05. **D**) Zika virus (ZIKV) seropositivity between febrile patients that were SARS-CoV-2 ELISA-positive versus SARS-CoV-2 ELISA-negative patients. Asterisk denotes p<0.05. Right: ZIKV IgG ELISA reactivity within both groups. Continuous line denotes the mean ELISA reactivity. N.s. not significant. **E**) ZIKV PRNT_50_ log_10_ results between 2019 febrile patients that were SARS-CoV-2 ELISA positive versus SARS-CoV-2 ELISA negative. Continuous line denotes the mean PRNT_50_ log_10_ reactivity **F**) ELISA ratio comparison between SARS-CoV-2 S1-IgA, S1-IgG, N-IgG ELISA and InBios SCoV-IgG ELISA positive or borderline patients with ZIKV-IgG ELISA. Dashed lines denote the ratio positivity threshold of >1.1 and borderline results between >0.9 to <1.1 defined by the manufacturer.

## Conclusion

We provide an assessment of SARS-CoV-2 antibody-based serologic diagnostics from Benin that reveals unspecific reactivity in up to 25% of febrile patients, possibly due to acute malaria. Limitations of our study include the small number of patients as well as limited patient metadata. Testing of sera for CMV and EBV by PCR may not have been sensitive due to lack of cell-associated viral nucleic acid, so that a potential impact of herpesvirus reactivation on serologic testing cannot be excluded. Additionally, we cannot exclude that dengue virus antibodies cross-reacted in the ZIKV ELISA in SARS-CoV-2 ELISA-positive patients (6). Nevertheless, our exhaustive analyses point at acute malaria as the most likely cause of the unspecific serologic reactivity observed, albeit other co-existing conditions also affecting testing cannot be excluded (6).

Unspecific reactivity in serologic tests might affect public health interventions in tropical regions, leading to an overestimate of SARS-CoV-2 circulation in regions where malaria is endemic and to misidentifications of SARS-CoV-2 hotspots. Additionally, target populations for vaccine campaigns once those become available might be missed, and coexistent diseases such as malaria might be overlooked based on false-positive SARS-CoV-2 results, leading to higher mortality from those endemic diseases (13, 14). The robustness of current and future SARS-CoV-2 serologic tests should be further assessed by multi-centric sero-epidemiologic studies from different tropical regions (15).

## Data Availability

Data available upon request

## Acknowledgments

We thank Arne Kühne, Wendy Jo-lei and Patricia Tscheak from the Institute of Virology, Charité, Berlin, Germany and Olfert Landt from Euroimmun GmbH, Germany.

This work was supported by the Deutsche Gesellschaft für Internationale Zusammenarbeit (GIZ) GmbH.

## Author Bio

Dr. Yadouleton is a medical entomologist in the Centre de Recherche Entomologique de Cotonou, Benin, head of the Laboratoire des Fièvres Hémorragiques in Cotonou, and a teacher at the University of Natitingou, Benin. His research interests include mosquito control and the diagnosis of viral hemorrhagic fevers.

## Notes

### Competing Interest Statement

The authors have declared no competing interest.

### Funding Statement

This work was supported by the Deutsche Gesellschaft fuer Internationale Zusammenarbeit (GIZ) GmbH.

### Author Declarations

This study was approved by the ethics committee of the Ministry of Health (Arrete 2020 No. 030/MS/DC/SGM/DNSP/CJ/SA/027SGG2020) and followed the Declaration of Helsinki. Written consent was obtained from all the patients participating in the study. Anonymized datasets were used, and all analysis of personally identifiable data took place only in the LFHB.

### Summary of Updates

A substantial peer review changed the manuscript to this updated version.

